# Benchmarking long-read variant sensitivity across ONT and PacBio platforms using known clinically reported variants in a cohort of critically ill newborns

**DOI:** 10.64898/2026.07.07.26357482

**Authors:** Colby T. Marvin, Joseph M. Devaney, Kati J. Buckingham, Jessica Noya, Kathryn M. Shively, Caitlin Jacques, Miranda Galey, Sophie H. Storz, Joy Goffena, April S. Berlyoung, Karynne E. Patterson, Tristan Shaffer, Christina Zakarian, Sean R. McGee, Joshua D. Smith, Lucas Lochovsky, Jonas A. Gustafson, Olivia M. Sommerland, Kailyn Anderson, Jamie Love-Nichols, Flavia M. Facio, Alexander V. Robertson, William J. Rowell, Juniper A. Lake, Andrew Carroll, Danny E. Miller, Chia L. Wei, Kirsty McWalter, Tara L. Wenger, University of Washington Center for Rare Disease Research, Britt Johnson, Michael J. Bamshad, Jessica X. Chong

## Abstract

Long-read whole genome sequencing (lrWGS) shows promise as an all-in-one test to detect clinically relevant variants and variants difficult to detect by current short-read whole genome sequencing (srWGS) pipelines. Comparisons between lrWGS and srWGS (or exome sequencing) pipelines will become commonplace as lrWGS is more widely adopted for clinical testing, particularly for individuals not diagnosed by srWGS. However, the sensitivity of lrWGS for detecting variants previously identified and prioritized by clinical srWGS has yet to be assessed. As part of the SeqFirst-neo study, a subset of critically ill newborns and their parents who underwent clinical srWGS also underwent lrWGS on the Oxford Nanopore Technologies (ONT) and Pacific Biosciences (PacBio) platforms. In total, 134 families were sequenced across multiple technologies including 128 families with clinical srWGS who were sequenced on both lrWGS platforms. We compared the variants reported by clinical testing with the variants identified by lrWGS. Among the 128 families sequenced on all three platforms, 89 SNV/indels and 14 SV/CNVs clinically reported by the srWGS testing pipeline were evaluated. All variants assessed in probands were ultimately detected by both lrWGS platforms, although three events were not detected prior to application of an updated variant caller, highlighting the rapid evolution of lrWGS variant calling. Additionally, breakpoint coordinates and event sizes often differed substantially between calls from srWGS and events called in lrWGS data. Our work demonstrates that while most clinically reported variants from srWGS can be detected by lrWGS pipelines, challenges remain when attempting direct comparisons, particularly for SV/CNVs.

## INTRODUCTION

Widespread adoption of whole exome sequencing and short-read whole genome sequencing (srWGS) has dramatically improved the diagnostic yield in pediatric patients with rare Mendelian conditions; however, these technologies are unable to detect epigenetic changes and have known limitations in capturing variants difficult to detect (VDD) such as complex SVs, repeat expansions, and variants in complex regions.^1–6^ In routine genetic testing workflows, this necessitates the use of complementary assays alongside srWGS to achieve a precise genetic diagnosis (PrGD) in a sizeable subset of patients.^7,8^

Long-read whole genome sequencing (lrWGS) is a promising new technology for its potential to capture both variants identified by srWGS and VDDs, theoretically eliminating the need for complementary assays and allowing for a single comprehensive clinical test.^9–12^ To date multiple studies have utilized lrWGS assays to identify VDDs particularly in cases where srWGS assays have failed to identify an explanatory (i.e., explains a person’s clinical findings) variant(s). However, the clinical implementation of lrWGS is still being investigated and a key first step is assessing the effectiveness of lrWGS pipelines to detect clinically relevant variants identified by existing clinical srWGS pipelines.^13–17^ While prior studies have investigated the concordance of variant calls produced from lrWGS and srWGS platforms in select samples, studies of concordance for clinically relevant variants in patient populations are limited.^18–21^

SeqFirst is a research initiative established to develop and test strategies to increase patient access to a PrGD in different pediatric care settings.^22^ In SeqFirst-neonatal (i.e., SeqFirst-neo) the aim is to improve access to a PrGD for infants with a critical illness by developing clinical workflows that support non-genetics providers, and facilitates greater access to genetic testing. We retrospectively performed lrWGS using both the Oxford Nanopore Technologies (ONT) and Pacific Biosciences (PacBio) platforms for families enrolled in SeqFirst-neo study who had received clinical srWGS. Herein we present a comparison of results across all three platforms.

## METHODS

### Study Cohort and Sequencing

Participants were enrolled in the SeqFirst-neo project which was reviewed and approved by the University of Washington Institutional Review Board (IRB #STUDY00008810). Application of exclusion criteria, study enrollment, and clinical rapid srWGS were conducted as previously described.^22^ A subset of 134 neonates and their parents for whom DNA of sufficient quality and quantity was available received lrWGS on a combination of ONT and PacBio platforms with 128 neonates sequenced on both lrWGS platforms in addition to clinical srWGS. The distribution of categorical admission diagnoses among these individuals was similar to the distribution observed in the full SeqFirst-neo cohort, with the majority (76/134 probands) of probands having either isolated or multiple congenital anomalies (Table S1).

Whole blood was collected from probands and parents for lrWGS and extracted using the New England Biolabs Monarch HMW DNA Extraction Kit for Cells & Blood. A Nanodrop Spectrophotometer ND-1000 was used to assess post extraction protein carryover. DNA samples sequenced on the ONT PromethION 24 instrument underwent library preparation with the ONT ligation kit (SQK-LSK110 or SQK-LSK114) and were sequenced in two batches: 62 probands and their parents (182 participants in total) were sequenced using the ONT R9.4.1 flow cell, 69 probands and their parents (180 participants in total) underwent mechanical shearing on the Diagenode Megaruptor 3 followed by fragment size analysis with an Agilent Femto Pulse and were sequenced using the ONT R10.4.1 flow cell. DNA samples sequenced on the PacBio Revio instrument first underwent mechanical shearing on either a Hamilton Microlab Prep or Diagenode Megaruptor 3 instrument followed by library preparation and sequencing using the PacBio SMRTbell prep kit 3.0 protocol, these included 132 probands and their parents (370 participants in total).

Sequencing on ONT was completed to 32.3x mean genome coverage (min-max 21.5-48.5x) on the R9.4.1 flow cell and 35.5x coverage (24.9– 47.3x) on the R10.4.1 flow cell. Sequencing on the PacBio Revio was completed to 31.1x coverage (6.5– 50.2x). Overall mean read lengths were: 19.6kb for ONT R9.4.1, 16.4kb for ONT R10.4.1, and 15.3kb for PacBio HiFi (Table S1).

Data generated on both the ONT and PacBio platforms were aligned to the reference human genome build hg38 (Figure 1). ONT data were basecalled using Dorado (v0.4.2) and aligned with minimap (v2.26)^23^, clair3 (v1.0.8)^24^ was used for single nucleotide variant and indel (SNV/indel) calling, sniffles2 (v2.3.3)^25^ was used for structural variant (SV) calling, hificnv (v1.0.0) was used for copy number variant (CNV) calling, and trgt (v1.5.1)^26^ was used for repeat expansion (RE) calling. PacBio data were basecalled using on-instrument SMRT Link software (v13.1) with DeepConsensus^27^ and aligned with pbmm2 (v1.10.0), DeepVariant (v1.4.0)^28^ was used for SNV/indel calling, pbsv (v2.9.0) was used for SV calling, hificnv (v.0.1.6) was used for CNV calling, and trgt (v1.0.0)^26^ was used for RE calling. On both datasets mitochondrial variants were called using the Mitoscope (v0.1.0) workflow with baldur^29^ variant calling and pb-CpG-tools (v3.0.0) followed by MethBat (v0.16.1) for calling methylation outliers. The Illumina srWGS data generated for clinical testing aligned to hg19 was realigned to hg38 with BWA-MEM (v0.7.15)^30^, SVs and CNVs were called using DRAGEN (v4.2.4)^31^ and Manta (v1.6.0)^32^.

**Figure 1.**
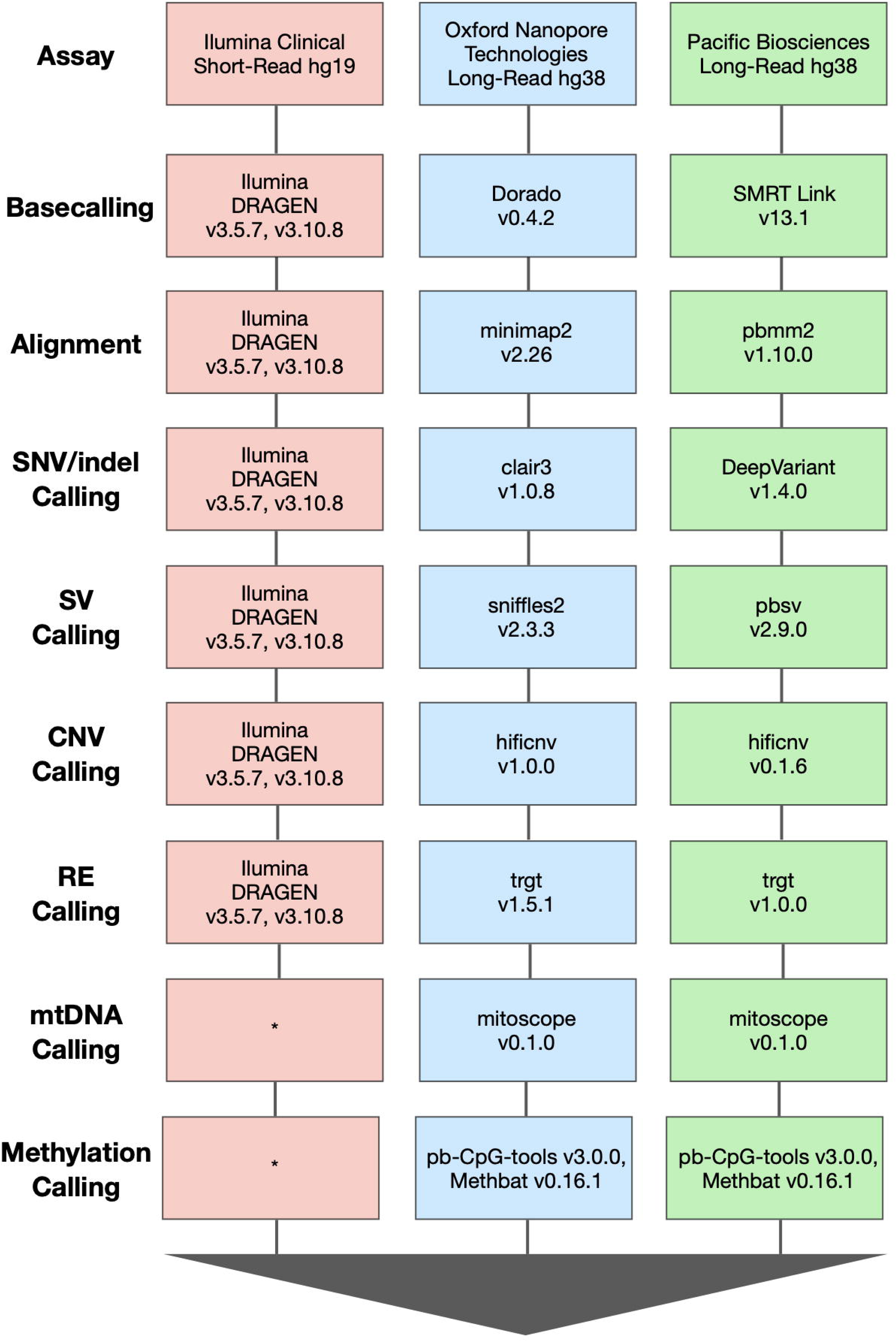
Data Processing Pipeline by Sequencing Technology Platform. Basecaller, alignment, and variant caller tools utilized for data generation across technology platforms. * Mitochondrial variants and methylation were called via separate tests ordered by providers in clinical short-read pipelines and were not called using srWGS data.

Following lrWGS data generation and srWGS data reprocessing, Peddy^33^ was used to confirm proband sex and familial relationships and identify samples with sequencing quality control (QC) metrics indicative of contamination. Data generated for one trio on the ONT R9.4.1 flow cell chemistry was subsequently excluded due to contamination.

After QC, 348 participants had genomes sequenced on each of the three platforms (clinical Illumina srWGS, ONT lrWGS, and PacBio lrWGS); 18 participants had data available from srWGS and PacBio lrWGS, seven participants had data from srWGS and ONT lrWGS, and four participants had data from ONT lrWGS and PacBio lrWGS (Figure 2). Family structures included 112 trios, 19 duos, and three singletons for a total of 377 participants (Table S1).

**Figure 2.**
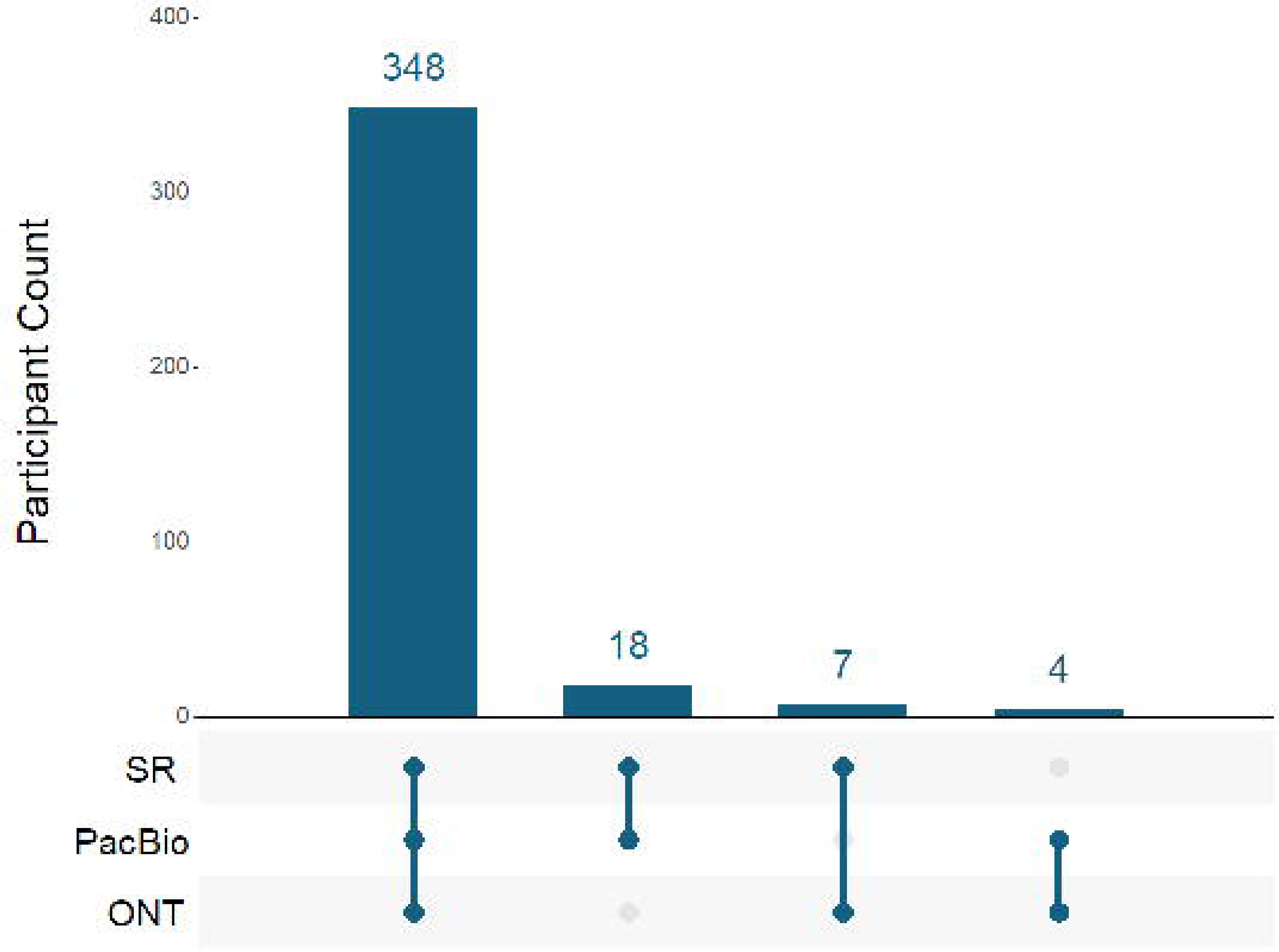
Participant Counts in SeqFirst-neo lrWGS Cohort by Sequencing Technology. Total counts of all participants (134 probands, 243 parents) from the SeqFirst-neo program selected for lrWGS grouped by the selection of technologies on which data was produced prior to data quality control including clinical rapid srWGS (SR), PacBio lrWGS (PacBio), and ONT lrWGS (ONT).

### Clinical Variant Ascertainment and Liftover

Evaluation of clinically reported variants was limited to the 128 probands with data generated on all three platforms. Clinical genetic test reports (GenomeXpress) issued by GeneDx were queried to pull all reported variants. Of probands sequenced on all three platforms, 74 probands had a total of 105 variants reported. Reported variants included 72 SNVs, 17 indels, six duplications, nine deletions, and one uniparental disomy (UPD) event. Two of these variants (chromosome 6 UPD and trisomy 21) were excluded from this study as the lrWGS variant calling workflows used in this study were not suited for their detection. All clinically reported variants had been called and reported using human genome reference build hg19 (Table S2, Table S3). American College of Medical Genetics and Genomics (ACMG) classifications of the 103 variants in our downstream analysis included 38 pathogenic, 21 likely pathogenic, and 44 variants of unknown significance (Figure 3).

**Figure 3.**
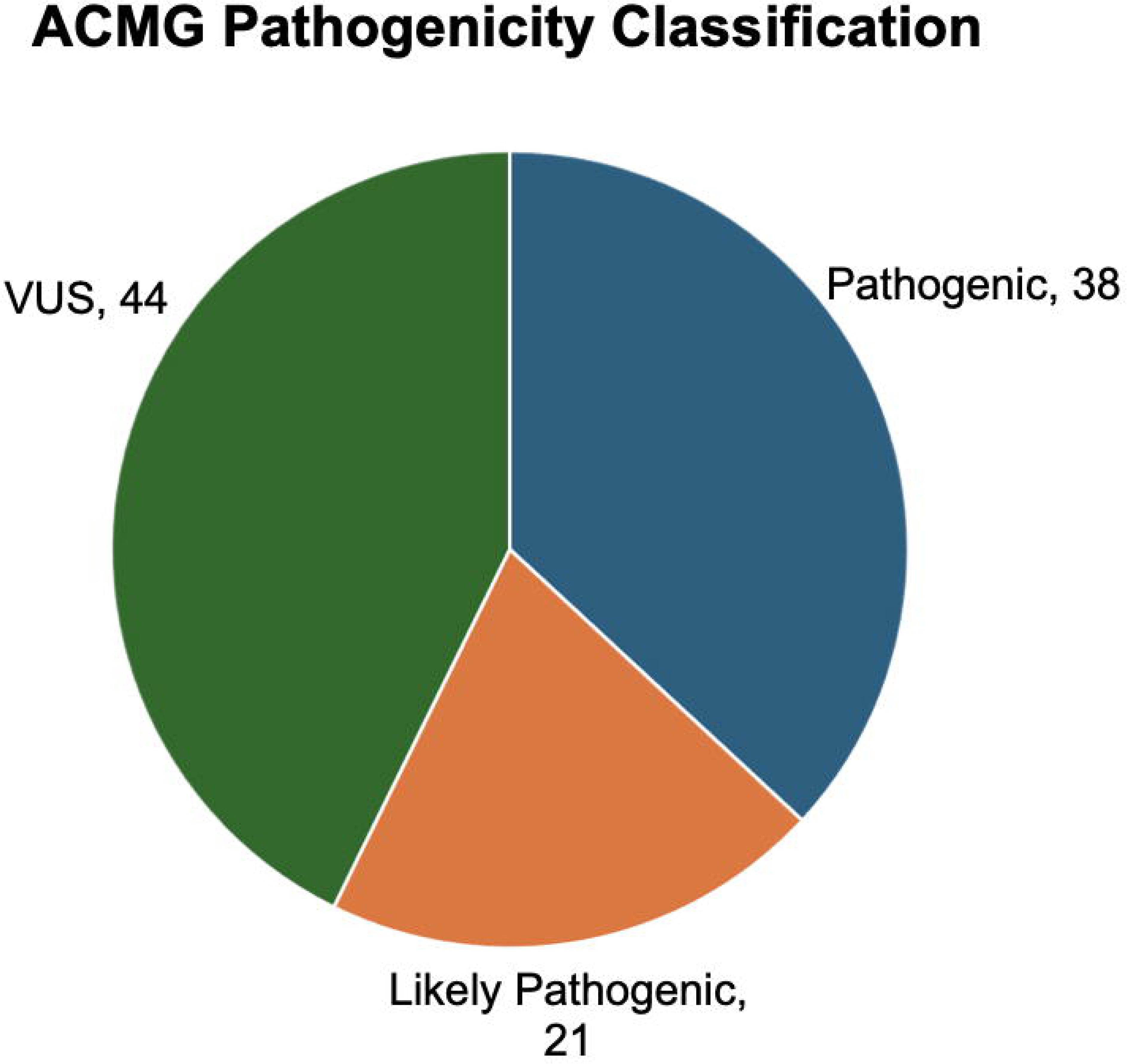
Clinically Reported Variants in SeqFirst-neo lrWGS cohort. ACMG pathogenicity classification of clinically reported variants in the SeqFirst-neo lrWGS cohort included in analysis.

Each clinically reported variant identified by srWGS on hg19 was lifted over to hg38 for comparison with lrWGS data. The UCSC LiftOver tool bulk liftover functionality was used for SNV/indels, while SV/CNV events were lifted over using both UCSC LiftOver and Broad LiftOver tools.^35,36^ Liftover of SV/CNV events was tested by providing the start and end coordinates as an interval (e.g., chr1:start-end) and by providing the positions independently (e.g. chr1:start and separately chr1:end) to the liftover tools.

### Variant Validation, Segregation, and Overlap Analysis

To assess the sensitivity of lrWGS pipelines for detecting clinically reported variants, we performed a series of queries on the lrWGS variant call file (VCF) outputs. These queries were categorized into two steps: Validation Check and Segregation Check. SV/CNV events underwent an additional step termed Overlap Check (Figure 4).

**Figure 4.**
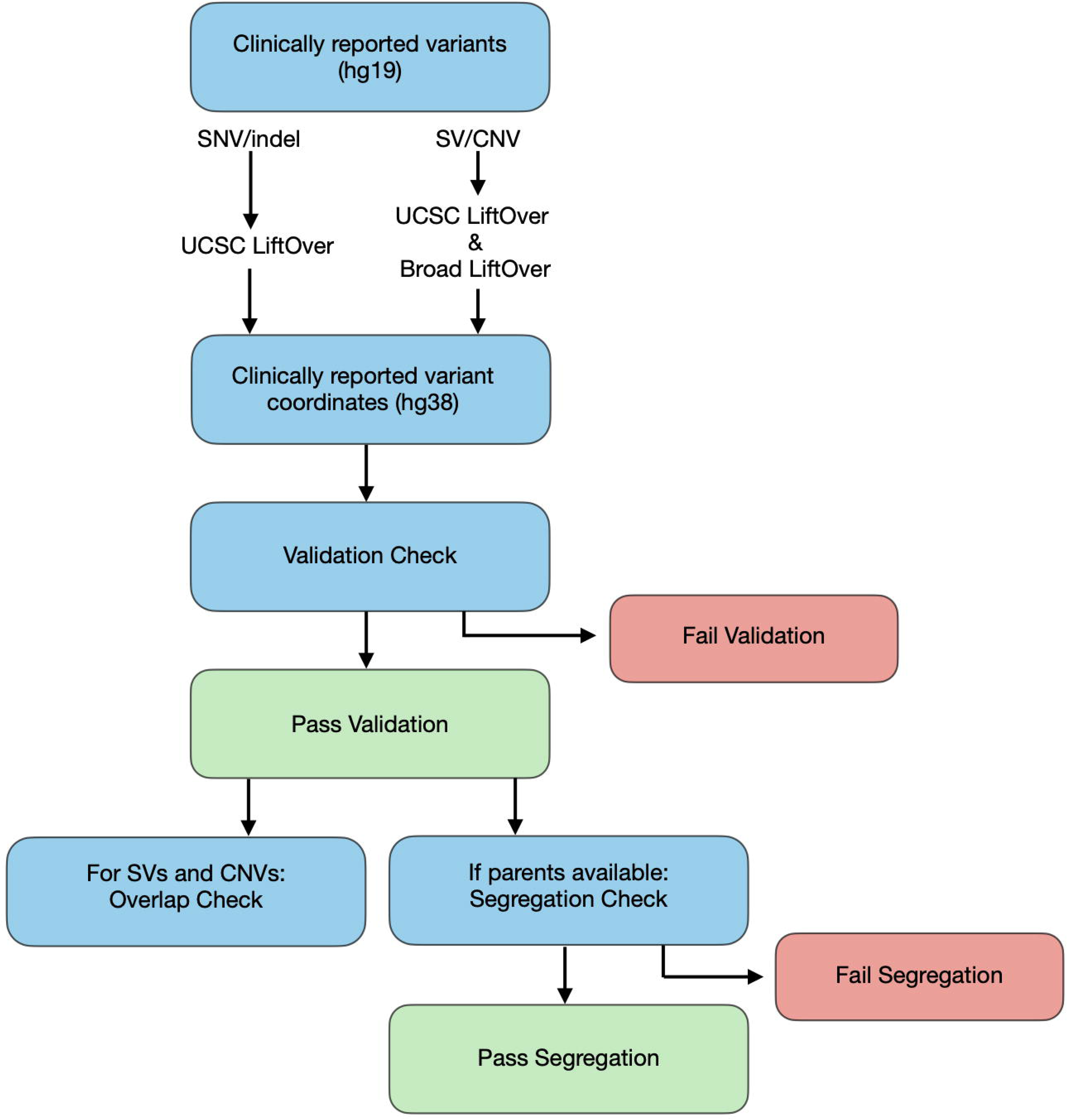
Analysis Workflow. Overview of analysis performed across lrWGS platforms of clinically reported variants assessed in the SeqFirst-neo lrWGS cohort.

SNV/indels were considered to pass Validation Check in a lrWGS platform if the same variant (same coordinate, reference, and alternate allele) was present in the lrWGS VCF at the hg38 liftover location with a ‘PASS’ filter flag. SV/CNVs were considered to pass Validation Check in a lrWGS platform if an event of the same type (i.e. duplication, deletion, translocation, inversion, etc.) and copy number was detected with a ‘PASS’ filter flag and intersected the coordinates of the reported event; the lrWGS event was also required to be no greater than 60% larger or smaller than the clinically reported event, as calculated by liftover to hg38 of the reported event’s coordinates, provided as an interval, using either liftover tool and subtracting the start from end coordinates.

Inherited variants were considered to pass Segregation Check in a lrWGS platform if the same variant was present in the parent indicated in the original srWGS report and it passed Validation Check. Variants clinically reported as *de novo* in srWGS were considered to pass Segregation Check only if both parental VCFs were available and neither detected the reported variant.

SV/CNV VCF outputs from the reprocessed srWGS data aligned to hg38 were queried to identify events of the same type and copy number with ‘PASS’ filter flags at the hg38 liftover locations using either tool corresponding to the clinically reported events on hg19.

Both liftover tools were used to calculate the hg38 coordinates of all SV/CNV variants that were originally called on hg19 and included in the clinical report. Then the regions for each SV/CNV passing Validation Check in the lrWGS data were intersected with both the hg38 liftover coordinates of the original hg19 variants as well as the coordinates of the variants that were called when the srWGS data were realigned and called on hg38. In the realigned srWGS data, if multiple separate events were called that spanned the clinically reported coordinates, the events were merged into a single region for purposes of overlap comparison. The minimum reciprocal overlap percentage between each pair of events was recorded. We termed this procedure Overlap Check.

## RESULTS

### SNV/indel Concordance

From the srWGS data a total 72 SNVs and 17 indels clinically reported on hg19 were lifted over to hg38 and queried for in proband and parent lrWGS caller VCF outputs from both ONT and PacBio platforms for Validation and Segregation Check (Figure 5A).

**Figure 5.**
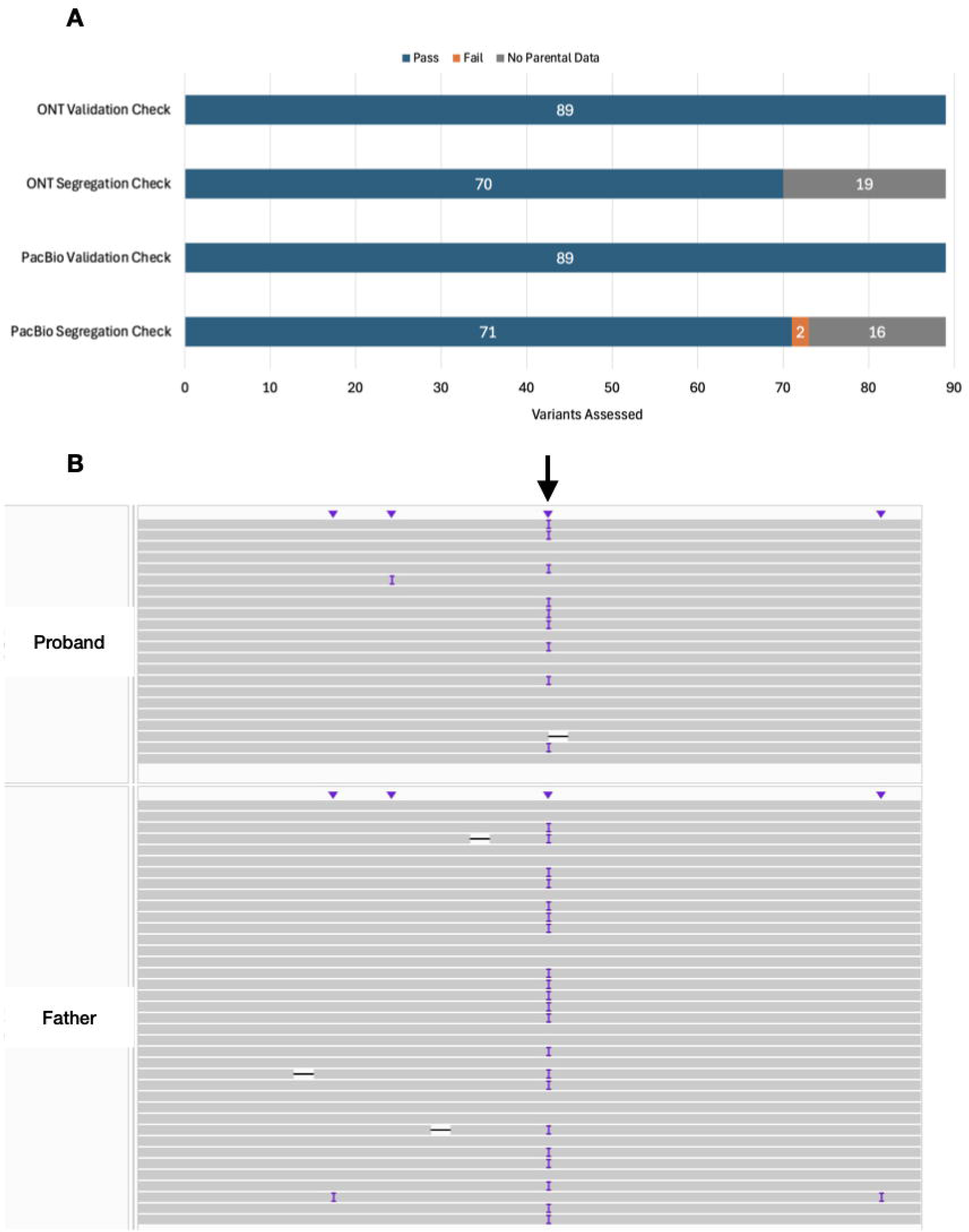
SNV/indel Validation and Segregation Check Results. (A) Validation and Segregation Check results of 89 clinically reported SNV/indels in the SeqFirst-neo lrWGS cohort split by lrWGS platform. (B) IGV overlay of 1bp insertion (arrow) in *ZFPM2* that failed initial Validation Check but was subsequently rescued after updating to DeepVariant v1.9.0.

All 89 SNV/indels passed Validation Check on the ONT platform. Segregation Check on the ONT platform was conducted for 70 out of 89 SNV/indels (55 SNVs, 15 indels) where parental data was available and successfully passed in all 70 instances (11 *de novo* confirmations, 59 inherited variant confirmations).

On the PacBio platform 88 out of 89 SNV/indels initially passed Validation Check. One paternally inherited 1bp insertion in *ZFPM2* failed Validation Check in sample 226 as it was present in the proband VCF (allele balance 0.41) but was genotyped as reference. Read coverage at the variant location was 22x and visual inspection in Integrated Genomics Viewer (IGV) did not identify any factors that could result in a missed call (Figure 5B). The 1bp indel was present and called with a ‘PASS’ GATK filter flag in the father’s VCF output (allele balance 0.56, 36x coverage at variant location). We subsequently updated DeepVariant from v1.4.0 to v1.9.0 and recalled this sample which successfully genotyped the variant in the proband, rescuing the single failing call for Validation Check. Segregation Check was then conducted over 73 out of 89 SNV/indels (58 SNVs, 15 indels) where parental data was available and successfully passed in 71 out of 73 instances (11 *de novo* confirmations, 60 inherited variant confirmations). In two instances Segregation Check failed in PacBio data; both were paternally inherited *PIEZO1* missense variants in a single father, sample 8, which had 14x mean genome coverage and read coverage of 3x and 5x over the variant sites.

### SV/CNV Concordance

A total of nine deletions and five duplications clinically reported on hg19 were lifted over to hg38 using both UCSC LiftOver and Broad LiftOver tools for Validation and Segregation Check (Figure 6). Deletions ranged in size from a 17.8kb event on chromosome 6 overlapping *TSC2* to a 58.2Mb event on chromosome X underlying Turner syndrome. Duplications ranged in size from a 549.9kb event on chr16 overlapping *CHAT* to a 10.2Mb event on chr12 overlapping multiple clinically relevant genes. For 12 out of 14 events, the UCSC LiftOver and Broad LiftOver tools returned nearly identical breakpoints (+/-1bp) that yielded the same total event size on hg38. However for two events both liftover tools returned an error when interval input was used resulting from the reported hg19 interval being split in hg38, a 6.2Mb deletion on chr15 in sample 103 and a 2.6Mb duplication on chr22 in sample 149. In both cases input of start and end positions independently to either liftover tool and combining the results into an interval resulted in a region that was much closer in size (5.7Mb for the chr15 deletion in sample 103 and 2.2Mb for the chr22 duplication in sample 149) to the clinically reported events on hg19. The position-derived liftover region from combining the liftover of the start and end positions was used for analysis for both previously errant events.

**Figure 6.**
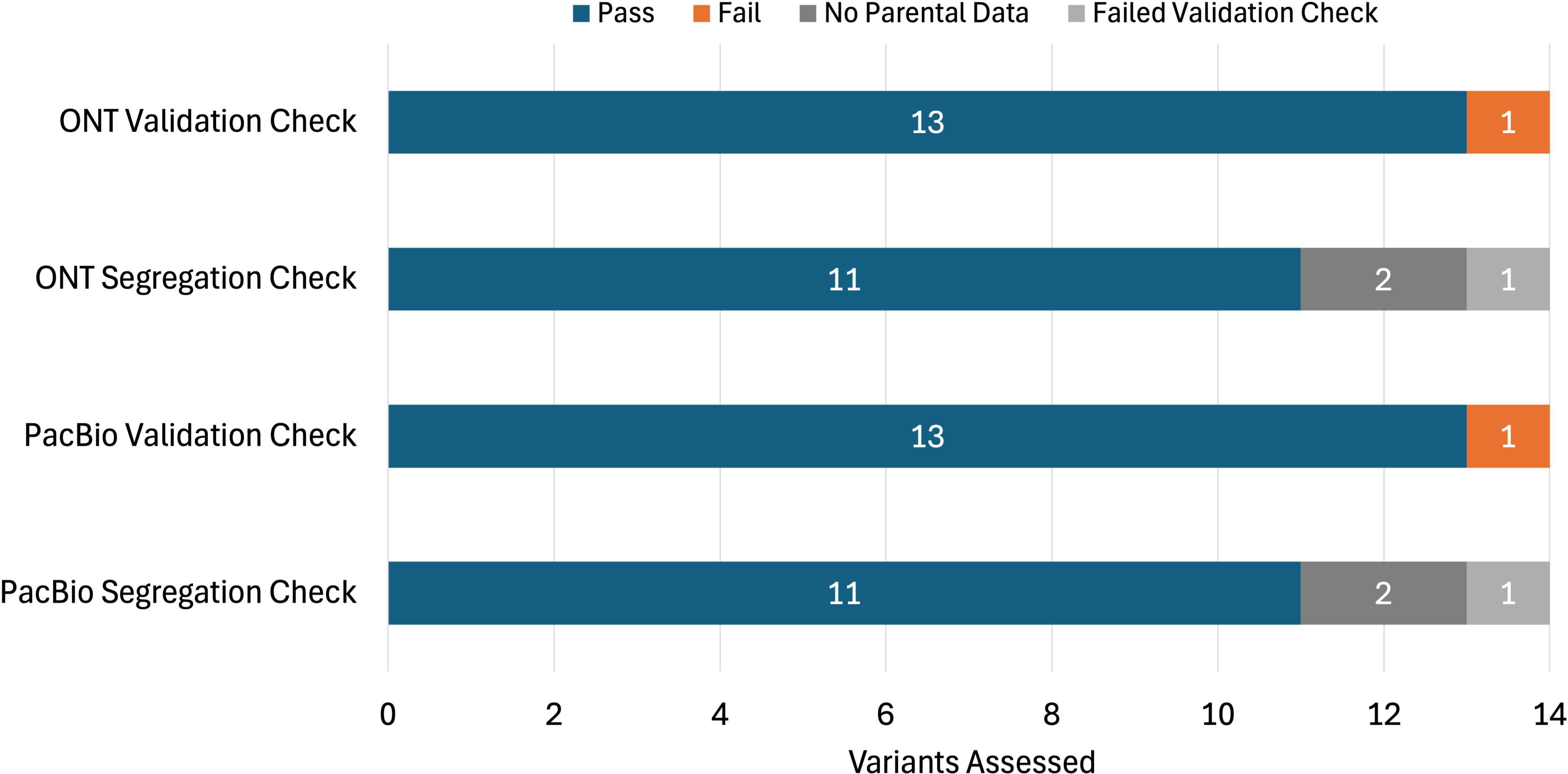
SV/CNV Validation and Segregation Check Results. Validation and Segregation Check results of 14 clinically reported SV/CNVs in the SeqFirst-neo lrWGS cohort split by lrWGS platform.

On the ONT platform, 12 out of 14 SV/CNV events initially passed Validation Check. The two remaining events, a 17.8kb deletion overlapping *TSC2* in sample 83 and a 52.4kb deletion overlapping *GLDC* in sample 336, were also the smallest events assessed and were not detected by either hificnv or sniffles2. We then used IGV to visually confirm that these deletions were present in the ONT data and recalled SVs for these two samples after updating sniffles2 from v2.3.3 to v2.6.2. This updated version of sniffles2 successfully rescued Validation Check for the 52.4kb deletion in sample 336. Notably, on inspection of the variant calls underlying the single remaining event in sample 83 failing Validation Check we identified a 41.7kb deletion in the ONT data that was 2.4 times larger than the clinically reported 17.8kb deletion. That these intermediate-sized SVs were missed by these tools was not surprising, versions of sniffles2 prior to v2.6.2 employed a filtering step which often flagged variants >10kb as false positives and filtered them out while hificnv is optimal for calling events larger than 100kb.^25^ Of the 12 events passing initial Validation Check, all were detected by hificnv. Parental data were available for 11 out of 13 events, and all 11 of these events passed Segregation Check.

On the PacBio platform, 13 out of 14 SV/CNV events passed initial Validation Check. Following inspection of the variant calls for the single event failing Validation Check in sample 83 we identified a 42.0kb deletion in the PacBio data 2.4 times larger than the clinically reported 17.8kb deletion, similar to the 41.7kb deletion identified by the ONT platform. This 42.0kb event and the second smallest event (52.4kb deletion overlapping *GLDC* in sample 336) passing Validation Check were successfully detected by pbsv, while the remaining 12 were successfully detected by hificnv. These detection differences are consistent with the design of the two callers (pbsv and hificnv) as pbsv is optimized for detection of deletions up to 100kb and duplications up to 20kb while hificnv is primarily designed to capture events larger than 100kb. All other SV/CNV events assessed exceeded the optimal detection size range for pbsv and fell within the target range for hificnv detection. Parental data was available for 11 out of 13 events passing Validation Check and all 11 of these passed Segregation Check.

We further investigated the single Validation Check failure remaining in both the ONT and PacBio datasets, the 41.7kb-42.0kb deletion detected in lrWGS data in sample 83 clinically reported as a 17.8kb deletion. We detected a 42.0kb deletion call (95.0-95.7% overlap to lrWGS calls) in both the original hg19 clinical srWGS callset and the hg38-recalled srWGS data. In collaboration with the original clinical sequencing laboratory, we determined that while the deletion in the hg19 clinical callset was 42kb, the coordinates and size provided in the clinical report for all SV/CNV events (14/14 assessed) were based on secondary orthogonal validation, in this case with an MRC Holland P337 *TSC2* Multiplex Ligation-dependent Probe Amplification (MLPA) assay, after which only the minimal supported region was reported. Subsequently a similar example was identified in sample 282 in which both lrWGS calls identified a larger 684-688kb deletion on chromosome 22 with lower overlap (77.0-77.5%) to the clinically reported region after liftover, but high overlap (95.9-96.4%) to the hg38-recalled srWGS data. Again, this discrepancy was a result of the clinically reported region being based on the secondary orthogonal validation, in this case an Agilent SurePrint MicroArray, in which a smaller 529.7kb deletion was identified.

These results followed from application of a liberal cut-off for defining a “matching” lrWGS event – we designated as “matching” any lrWGS call that overlapped at least 1bp of the clinically reported event interval (after liftover) and was within +/-60% of the size of the clinically reported event. The use of a more stringent matching cutoff, such as requiring the lrWGS call to be +/-10% of the size of the clinically reported event results in the majority of lrWGS events failing to match against the clinically reported event (6/14 matches on ONT, 7/14 matches on PacBio). This drop-off in matching is, in part, a result of using the coordinates from the clinical report, which were the product of the original diagnostic laboratory’s policies for reporting the minimally supported region after orthogonal validation. Accordingly, comparing the lrWGS calls against the coordinates and sizes of the raw hg19 srWGS event calls after liftover to hg38 with a +/-60% cutoff yields matches for all 14 events assessed across both lrWGS platforms, while the more stringent +/-10% cutoff yields 7/14 matches on ONT and 9/14 matches on PacBio.

Finally, we systematically assessed the minimum reciprocal overlap between the coordinate pair for each lrWGS event and two sets of coordinates for the same events derived from the srWGS data—liftover of the clinically reported coordinate pairs to hg38 and hg38-native coordinate pairs from the hg38-realigned srWGS (Figure 7A). Of 13 events passing Validation Check on both lrWGS platforms, the percent overlap between the events in the ONT callset and the coordinates of the clinically reported events after hg38 liftover or hg38 realignment ranged from 63.0-100.0% with mean overlap 86.5% for both Broad LiftOver and UCSC LiftOver. Overlap with calls from the hg38-realigned srWGS data ranged from 56.0-100.0% with mean overlap 88.9% (Figure 7A). Overlap between the 13 events passing Validation Check in the PacBio callset and the coordinates of the clinically reported events after hg38 liftover ranged from 64.9-100.0% with mean overlap 87.1% for both Broad LiftOver and UCSC LiftOver. Overlap with calls from the hg38-realigned srWGS data ranged from 57.7-100.0% with mean overlap 87.8% (Figure 7A). We also assessed reciprocal overlap between PacBio and ONT SV/CNV calls, which was 79.5-100.0% for all 14 events assessed.

**Figure 7.**
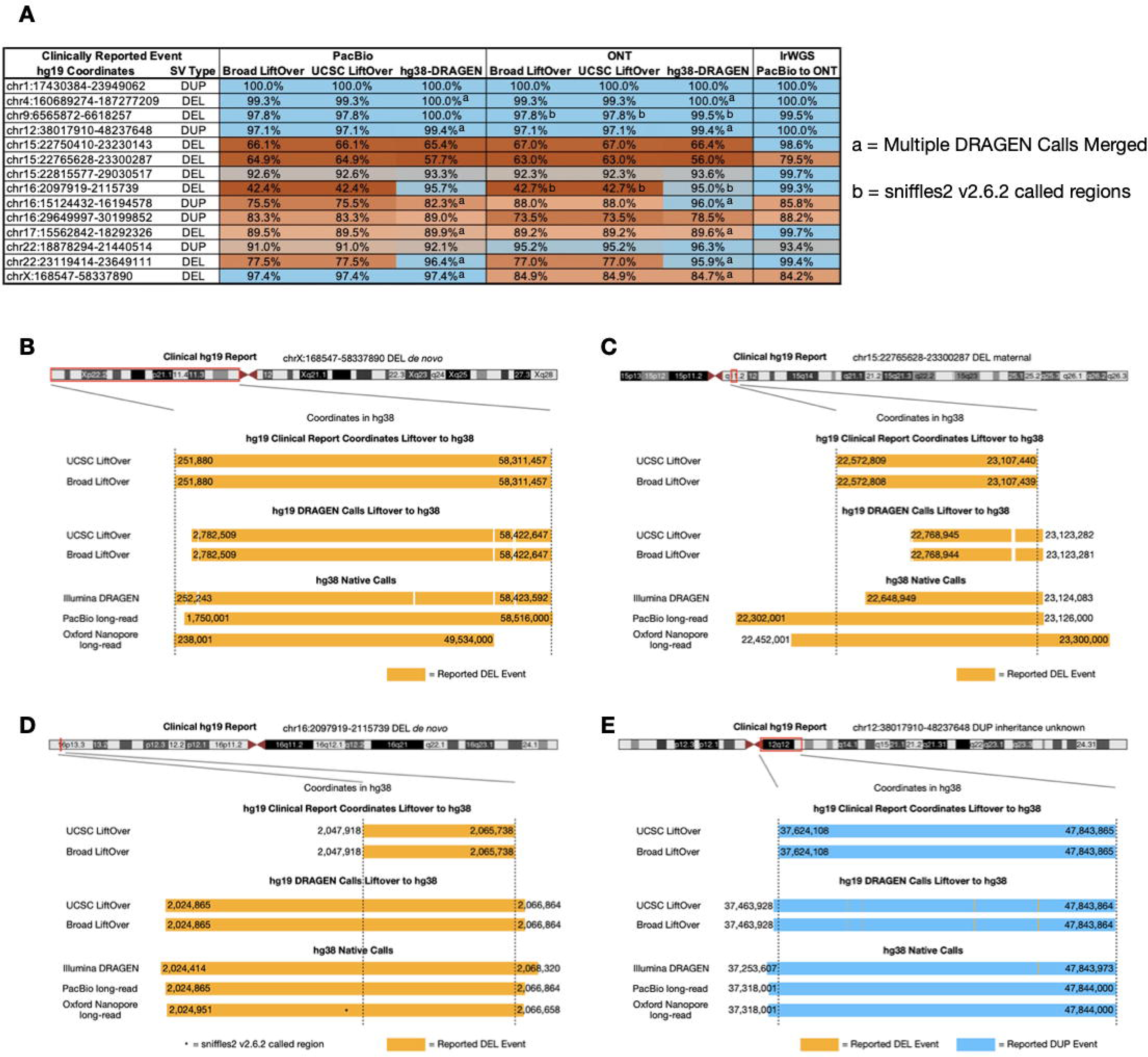
Overlap Check Results. (A) Overlap of lrWGS called events compared to hg19 clinically reported events after liftover and hg38 srWGS called events. Example overlap comparisons are shown including (B) event with substantial (1.5, 9.0Mb) breakpoint call differences between lrWGS platforms, (C) event with low overlap (56.0-64.9%) between lrWGS and srWGS call regions, (D) event in which srWGS and both lrWGS platforms called a region 2.4 times larger than what was clinically reported after liftover to hg38, and (E) event with high (97.1-100%) overlap between the clinically reported region after liftover to hg38, srWGS call region, and lrWGS call regions.

## DISCUSSION

ONT and PacBio lrWGS were used successfully to detect all the SNV/indels and SV/CNVs that were reported by clinical srWGS testing. Where parental data were available, we confirmed segregation in all but two indels on PacBio and both segregation discrepancies were explained by low coverage in the parental samples. These samples likely would have been sequenced to higher coverage if the lrWGS was generated in a clinical setting.^37^ The SNV/indel on PacBio and two SV/CNVs on ONT that were initially missed were subsequently rescued using updated variant callers. This highlights the rapidly evolving landscape of lrWGS technologies compared to the analysis of srWGS data generation and calling.

Our experience attempting to confirm the presence/absence in the hg38-aligned lrWGS of SV/CNVs that were clinically reported on hg19 highlights additional challenges to routine implementation. Notably, after our original analysis was completed, UCSC LiftOver updated their web interface to no longer accept interval input for hg19 regions which would map to multiple hg38 intervals and instead directs users to use single position inputs. Prior to this update, use of interval input for liftover of these regions (2/14 assessed) returned multiple regions with no corresponding overlap to single position input results from the Broad LiftOver tool. This is a known challenge for using liftover between genome assemblies as intervals spanning a genomic region that is not contiguous in the target genome assembly may end up mapped to multiple segments of smaller size and location.^38,39^

For each clinically reported SV/CNV, we found substantial variation in extent of overlap between the original clinically reported hg19 call when lifted over to hg38 and the “matching” hg38-native event in the lrWGS data (63-100%) as well as between the same event called by the two lrWGS pipelines (79.5-100%). This substantial variation in the extent of overlap / length and coordinates (Figures 7B, 7C) of SV/CNVs suggests that laboratories and providers performing lrWGS on patients with prior clinical srWGS test results on hg19, or even patients who received lrWGS on a different sequencing platform, should anticipate the possibility of substantial differences in event coordinates and sizes. Similar challenges are present when comparing SV/CNVs identified by chromosomal microarray (CMA) or older technologies to events detected by short read technologies (panels, ES, srWGS), but we observed larger differences in event breakpoint coordinates (55bp-8.7Mb difference in called events) than previously observed for CMA vs. srWGS.^40,41^ These differences could lead to discrepant ACMG pathogenicity classifications and clinical relevance assessments.

In addition to technical differences (e.g. liftover, calling algorithms, and sequencing platforms), we found that an event’s coordinates in the clinical report may not be the direct product of the original variant calls, but may instead be based on multiple sources including: the coordinates identified by orthogonal validation methods (Figure 7D), the smallest overlapping set of coordinates between a validation technology and the original srWGS call, the coordinates of the clinically relevant region as determined by manual review, or the original srWGS event coordinates. This complicates efforts to compare newer SV/CNV calls directly against prior clinically reported events. A more comprehensive assessment will be necessary to establish guidelines for assessing matching between hg19 SV/CNVs (likely to be common in prior clinical reports for microarray, exome, and srWGS tests) lifted over to hg38 and hg38-native SV/CNV calls, especially if the latter were called from a lrWGS-based pipeline.

In summary, our results show that both the ONT and PacBio lrWGS pipelines were able to detect all SNV/indels and SV/CNVs in proband samples for clinically reported variants that were previously reported by clinical srWGS testing, but the overlap in size and position of some SV/CNV events varied substantially. This result is consistent with prior reports of high concordance between lrWGS and srWGS variant calls based on sequencing of validation samples such as Genome in a Bottle.^42^ However, our focus on variants that were previously selected for inclusion in a clinical genetic testing report allowed us to confirm high sensitivity for these clinically relevant variants. That three variants were initially missed using older versions of lrWGS variant callers and rescued after updating to newer versions highlights the rapidly evolving landscape of lrWGS analysis tools and presents a possible limitation to widespread adoption for this technology for clinical testing in the near future. Finally, our results highlight practical considerations to address as future clinical applications of lrWGS-based genetic testing emerge alongside ongoing srWGS-based testing.

## Data and code availability

All data regarding reported variants and presence in lrWGS outputs is available in the supplemental information. The srWGS, ONT lrWGS, and PacBio lrWGS data have been deposited and are available in NHGRI’s Analysis Visualization and Informatics Lab-space (AnVIL) (accession no. phs003047, “internal_project_id” contains SeqFirst-neo).

## Data Availability

All data produced are available in the NHGRI Analysis Visualization and Informatics Lab-space (AnVIL) (accession no. phs003047 internal_project_id contains SeqFirst-neo).

## Acknowledgments

We thank the families for their participation and support. We thank the faculty and staff of the SCH NICU and the CICU. We would like to thank Evan E. Eichler, an investigator of the Howard Hughes Medical Institute, for support with initial project planning, setup, and sequencing. Financial support was provided by grants from GeneDx, the Brotman-Baty Institute, and University of Washington Center for Rare Disease Research (UW-CRDR), with additional contributions from NHGRI grants U01 HG011744, UM1 HG006493, and U24 HG011746. DEM is supported by NIH grant DP5OD033357. Sequencing data generated on the Oxford Nanopore Technologies platform was in part supported by University of Washington Institutional Review Board #STUDY00008810. The content is solely the responsibility of the authors and does not necessarily represent the official views of the National Institutes of Health.

## Web Resources

Dorado: https://github.com/nanoporetech/dorado

minimap: https://github.com/lh3/minimap2

clair3: https://github.com/HKU-BAL/clair3

sniffles2: https://github.com/fritzsedlazeck/sniffles

hificnv: https://github.com/PacificBiosciences/HiFiCNV

trgt: https://github.com/pacificBiosciences/trgt/

pbmm2: https://github.com/PacificBiosciences/pbmm2

DeepVariant: https://github.com/google/deepvariant

pbsv: https://github.com/PacificBiosciences/pbsv

mitoscope: https://github.com/czakarian/mitoscope

pb-CpG-tools: https://github.com/PacificBiosciences/pb-CpG-tools

Methbat: https://github.com/PacificBiosciences/MethBat

BWA-MEM: https://github.com/lh3/BWA

Manta: https://github.com/Illumina/manta

Peddy: https://github.com/brentp/peddy

UCSC LiftOver: https://genome.ucsc.edu/cgi-bin/hgLiftOver

Broad LiftOver: https://liftover.broadinstitute.org/

## References

1. Bamshad, M.J., Nickerson, D.A., and Chong, J.X. (2019). Mendelian Gene Discovery: Fast and Furious with No End in Sight. The American Journal of Human Genetics 105, 448–455. 10.1016/j.ajhg.2019.07.011.

2. Chong, J.X., Buckingham, K.J., Jhangiani, S.N., Boehm, C., Sobreira, N., Smith, J.D., Harrell, T.M., McMillin, M.J., Wiszniewski, W., Gambin, T., et al. (2015). The Genetic Basis of Mendelian Phenotypes: Discoveries, Challenges, and Opportunities. The American Journal of Human Genetics 97, 199–215. 10.1016/j.ajhg.2015.06.009.

3. Tan, N.B., Tan, T.Y., Martyn, M.M., Savarirayan, R., Amor, D.J., Moody, A., White, S.M., and Stark, Z. (2019). Diagnostic and service impact of genomic testing technologies in a neonatal intensive care unit. Journal of Paediatrics and Child Health 55, 1309–1314. 10.1111/jpc.14398.

4. Lee, H.-F., Chi, C.-S., and Tsai, C.-R. (2021). Diagnostic yield and treatment impact of whole-genome sequencing in paediatric neurological disorders. Developmental Medicine & Child Neurology 63, 934–938. 10.1111/dmcn.14722.

5. Lindstrand, A., Eisfeldt, J., Pettersson, M., Carvalho, C.M.B., Kvarnung, M., Grigelioniene, G., Anderlid, B.-M., Bjerin, O., Gustavsson, P., Hammarsjö, A., et al. (2019). From cytogenetics to cytogenomics: whole-genome sequencing as a first-line test comprehensively captures the diverse spectrum of disease-causing genetic variation underlying intellectual disability. Genome Med 11, 68. 10.1186/s13073-019-0675-1.

6. Kernohan, K.D., and Boycott, K.M. (2024). The expanding diagnostic toolbox for rare genetic diseases. Nat Rev Genet 25, 401–415. 10.1038/s41576-023-00683-w.

7. Hu, L., Ru, K., Zhang, L., Huang, Y., Zhu, X., Liu, H., Zetterberg, A., Cheng, T., and Miao, W. (2014). Fluorescence in situ hybridization (FISH): an increasingly demanded tool for biomarker research and personalized medicine. Biomark Res 2, 3. 10.1186/2050-7771-2-3.

8. Yang, Y., del Gaudio, D., Santani, A., and Scott, S.A. (2024). Applications of genome sequencing as a single platform for clinical constitutional genetic testing. Genetics in Medicine Open 2, 101840. 10.1016/j.gimo.2024.101840.

9. Miller, D.E., Sulovari, A., Wang, T., Loucks, H., Hoekzema, K., Munson, K.M., Lewis, A.P., Fuerte, E.P.A., Paschal, C.R., Walsh, T., et al. (2021). Targeted long-read sequencing identifies missing disease-causing variation. The American Journal of Human Genetics 108, 1436–1449. 10.1016/j.ajhg.2021.06.006.

10. Mastrorosa, F.K., Miller, D.E., and Eichler, E.E. (2023). Applications of long-read sequencing to Mendelian genetics. Genome Med 15, 42. 10.1186/s13073-023-01194-3.

11. Logsdon, G.A., Vollger, M.R., and Eichler, E.E. (2020). Long-read human genome sequencing and its applications. Nat Rev Genet 21, 597–614. 10.1038/s41576-020-0236-x.

12. Del Gobbo, G.F., and Boycott, K.M. (2025). The additional diagnostic yield of long-read sequencing in undiagnosed rare diseases. Genome Res 35, 559–571. 10.1101/gr.279970.124.

13. Dawood, M., Heavner, B., Wheeler, M.M., Ungar, R.A., LoTempio, J., Wiel, L., Berger, S., Bernstein, J.A., Chong, J.X., Délot, E.C., et al. (2025). GREGoR: accelerating genomics for rare diseases. Nature 647, 331–342. 10.1038/s41586-025-09613-8.

14. Hiatt, S.M., Lawlor, J.M.J., Handley, L.H., Latner, D.R., Bonnstetter, Z.T., Finnila, C.R., Thompson, M.L., Boston, L.B., Williams, M., Rodriguez Nunez, I., et al. (2024). Long-read genome sequencing and variant reanalysis increase diagnostic yield in neurodevelopmental disorders. Genome Res 34, 1747–1762. 10.1101/gr.279227.124.

15. Miller, D.E., Lee, L., Galey, M., Kandhaya-Pillai, R., Tischkowitz, M., Amalnath, D., Vithlani, A., Yokote, K., Kato, H., Maezawa, Y., et al. (2022). Targeted long-read sequencing identifies missing pathogenic variants in unsolved Werner syndrome cases. J Med Genet 59, 1087–1094. 10.1136/jmedgenet-2022-108485.

16. Eisfeldt, J., Ek, M., Nordenskjöld, M., and Lindstrand, A. (2025). Toward clinical long-read genome sequencing for rare diseases. Nat Genet 57, 1334–1343. 10.1038/s41588-025-02160-y.

17. Thiffault, I., Farrow, E., Barrett, C., Scott, M., Ross, A., Means, J.C., Cheung, W.A., Johnson, A.F., Koseva, B., McLennan, R., et al. (2025). Clinical Long-Read Sequencing Test for Genetic Disease Diagnosis. JAMA Pediatr 179, 1355–1357. 10.1001/jamapediatrics.2025.3320.

18. Chaisson, M.J.P., Sanders, A.D., Zhao, X., Malhotra, A., Porubsky, D., Rausch, T., Gardner, E.J., Rodriguez, O.L., Guo, L., Collins, R.L., et al. (2019). Multi-platform discovery of haplotype-resolved structural variation in human genomes. Nat Commun 10, 1784. 10.1038/s41467-018-08148-z.

19. Sen, S., Handler, H.P., Victorsen, A., Flaten, Z., Ellison, A., Knutson, T.P., Munro, S.A., Martinez, R.J., Billington, C.J., Laffin, J.J., et al. (2025). Validation of a comprehensive long-read sequencing platform for broad clinical genetic diagnosis. Front Genet 16, 1499456. 10.3389/fgene.2025.1499456.

20. Zhao, X., Collins, R.L., Lee, W.-P., Weber, A.M., Jun, Y., Zhu, Q., Weisburd, B., Huang, Y., Audano, P.A., Wang, H., et al. (2021). Expectations and blind spots for structural variation detection from long-read assemblies and short-read genome sequencing technologies. Am J Hum Genet 108, 919–928. 10.1016/j.ajhg.2021.03.014.

21. Wagner, J., Olson, N.D., Harris, L., Khan, Z., Farek, J., Mahmoud, M., Stankovic, A., Kovacevic, V., Yoo, B., Miller, N., et al. (2022). Benchmarking challenging small variants with linked and long reads. Cell Genom 2, 100128. 10.1016/j.xgen.2022.100128.

22. Wenger, T.L., Scott, A., Kruidenier, L., Sikes, M., Keefe, A., Buckingham, K.J., Marvin, C.T., Shively, K.M., Bacus, T., Sommerland, O.M., et al. (2025). SeqFirst: Building equity access to a precise genetic diagnosis in critically ill newborns. Am J Hum Genet 112, 508–522. 10.1016/j.ajhg.2025.02.003.

23. Li, H. (2018). Minimap2: pairwise alignment for nucleotide sequences. Bioinformatics 34, 3094–3100. 10.1093/bioinformatics/bty191.

24. Zheng, Z., Li, S., Su, J., Leung, A.W.-S., Lam, T.-W., and Luo, R. (2022). Symphonizing pileup and full-alignment for deep learning-based long-read variant calling. Nat Comput Sci 2, 797–803. 10.1038/s43588-022-00387-x.

25. Smolka, M., Paulin, L.F., Grochowski, C.M., Horner, D.W., Mahmoud, M., Behera, S., Kalef-Ezra, E., Gandhi, M., Hong, K., Pehlivan, D., et al. (2024). Detection of mosaic and population-level structural variants with Sniffles2. Nat Biotechnol 42, 1571–1580. 10.1038/s41587-023-02024-y.

26. Dolzhenko, E., English, A., Dashnow, H., De Sena Brandine, G., Mokveld, T., Rowell, W.J., Karniski, C., Kronenberg, Z., Danzi, M.C., Cheung, W.A., et al. (2024). Characterization and visualization of tandem repeats at genome scale. Nat Biotechnol 42, 1606–1614. 10.1038/s41587-023-02057-3.

27. Baid, G., Cook, D.E., Shafin, K., Yun, T., Llinares-López, F., Berthet, Q., Belyaeva, A., Töpfer, A., Wenger, A.M., Rowell, W.J., et al. (2023). DeepConsensus improves the accuracy of sequences with a gap-aware sequence transformer. Nat Biotechnol 41, 232–238. 10.1038/s41587-022-01435-7.

28. Poplin, R., Chang, P.-C., Alexander, D., Schwartz, S., Colthurst, T., Ku, A., Newburger, D., Dijamco, J., Nguyen, N., Afshar, P.T., et al. (2018). A universal SNP and small-indel variant caller using deep neural networks. Nat Biotechnol 36, 983–987. 10.1038/nbt.4235.

29. Keraite, I., Becker, P., Canevazzi, D., Frias-López, C., Dabad, M., Tonda-Hernandez, R., Paramonov, I., Ingham, M.J., Brun-Heath, I., Leno, J., et al. (2022). A method for multiplexed full-length single-molecule sequencing of the human mitochondrial genome. Nat Commun 13, 5902. 10.1038/s41467-022-33530-3.

30. Li, H. (2013). Aligning sequence reads, clone sequences and assembly contigs with BWA-MEM. Preprint at arXiv, 10.48550/arXiv.1303.3997 https://doi.org/10.48550/arXiv.1303.3997.

31. Behera, S., Catreux, S., Rossi, M., Truong, S., Huang, Z., Ruehle, M., Visvanath, A., Parnaby, G., Roddey, C., Onuchic, V., et al. (2025). Comprehensive genome analysis and variant detection at scale using DRAGEN. Nat Biotechnol 43, 1177–1191. 10.1038/s41587-024-02382-1.

32. Chen, X., Schulz-Trieglaff, O., Shaw, R., Barnes, B., Schlesinger, F., Källberg, M., Cox, A.J., Kruglyak, S., and Saunders, C.T. (2016). Manta: rapid detection of structural variants and indels for germline and cancer sequencing applications. Bioinformatics 32, 1220–1222. 10.1093/bioinformatics/btv710.

33. Pedersen, B.S., and Quinlan, A.R. (2017). Who’s Who? Detecting and Resolving Sample Anomalies in Human DNA Sequencing Studies with Peddy. The American Journal of Human Genetics 100, 406–413. 10.1016/j.ajhg.2017.01.017.

34. Green, R.C., Berg, J.S., Grody, W.W., Kalia, S.S., Korf, B.R., Martin, C.L., McGuire, A.L., Nussbaum, R.L., O’Daniel, J.M., Ormond, K.E., et al. (2013). ACMG recommendations for reporting of incidental findings in clinical exome and genome sequencing. Genet Med 15, 565–574. 10.1038/gim.2013.73.

35. Hinrichs, A.S., Karolchik, D., Baertsch, R., Barber, G.P., Bejerano, G., Clawson, H., Diekhans, M., Furey, T.S., Harte, R.A., Hsu, F., et al. (2006). The UCSC Genome Browser Database: update 2006. Nucleic Acids Res 34, D590–D598. 10.1093/nar/gkj144.

36. Genovese, G., Rockweiler, N.B., Gorman, B.R., Bigdeli, T.B., Pato, M.T., Pato, C.N., Ichihara, K., and McCarroll, S.A. (2024). BCFtools/liftover: an accurate and comprehensive tool to convert genetic variants across genome assemblies. Bioinformatics 40, btae038. 10.1093/bioinformatics/btae038.

37. Devaney, J.M., Chong, J.X., Lopes, P.C., Noya, J., Berlyoung, A.S., Yusuff, S., Lynch, S., Brandon, R., Hruska, K.S., Lochovsky, L., et al. (2026). Sensitivity of HiFi long-read genome sequencing for difficult-to-detect pathogenic variants when applied to real-world clinical laboratory samples. The American Journal of Human Genetics 0. 10.1016/j.ajhg.2026.04.001.

38. Gao, B., Huang, Q., and Baudis, M. (2018). segment_liftover : a Python tool to convert segments between genome assemblies. F1000Res *7*, 319. 10.12688/f1000research.14148.2.

39. Luu, P.-L., Ong, P.-T., Dinh, T.-P., and Clark, S.J. (2020). Benchmark study comparing liftover tools for genome conversion of epigenome sequencing data. NAR Genom Bioinform 2, lqaa054. 10.1093/nargab/lqaa054.

40. Gross, A.M., Ajay, S.S., Rajan, V., Brown, C., Bluske, K., Burns, N.J., Chawla, A., Coffey, A.J., Malhotra, A., Scocchia, A., et al. (2019). Copy-number variants in clinical genome sequencing: deployment and interpretation for rare and undiagnosed disease. Genetics in Medicine 21, 1121–1130. 10.1038/s41436-018-0295-y.

41. Stavropoulos, D.J., Merico, D., Jobling, R., Bowdin, S., Monfared, N., Thiruvahindrapuram, B., Nalpathamkalam, T., Pellecchia, G., Yuen, R.K.C., Szego, M.J., et al. (2016). Whole-genome sequencing expands diagnostic utility and improves clinical management in paediatric medicine. npj Genomic Med 1, 15012. 10.1038/npjgenmed.2015.12.

42. Olson, N.D., Wagner, J., McDaniel, J., Stephens, S.H., Westreich, S.T., Prasanna, A.G., Johanson, E., Boja, E., Maier, E.J., Serang, O., et al. (2022). PrecisionFDA Truth Challenge V2: Calling variants from short and long reads in difficult-to-map regions. Cell Genomics 2, 100129. 10.1016/j.xgen.2022.100129.

